# A Model-agnostic Computational Method for Discovering Gene–Phenotype Relationships and Inferring Gene Networks via *in silico* Gene Perturbation

**DOI:** 10.1101/2024.02.21.24303141

**Authors:** Rastko Stojšin, Xiangning Chen, Zhongming Zhao

**Affiliations:** Center for Precision Health, McWilliams School of Biomedical Informatics, The University of Texas Health Science Center at Houston, Houston, TX 77030, USA; MD Anderson Cancer Center UTHealth Graduate School of Biomedical Sciences, Houston, TX 77030, USA

**Author notes:** To whom correspondence should be addressed: Zhongming Zhao, Ph.D. Center for Precision Health, McWilliams School of Biomedical Informatics, The University of Texas Health Science Center at Houston 7000 Fannin St., Suite 600, Houston, TX 77030, Xiangning Chen, Ph.D. Center for Precision Health, McWilliams School of Biomedical Informatics, The University of Texas Health Science Center at Houston 7000 Fannin St., Suite 600, Houston, TX 77030. Email addresses: RS XC ZZ.

**Keywords:** deep learning, genotype–phenotype mapping, biomarker discovery, gene networks, bioinformatics

## Abstract

**Background:** Deep learning architectures have advanced genotype‒phenotype mappings with precision but often obscure the roles of specific genes and their interactions. Our research introduces a model-agnostic computational methodology, capitalizing on the analytical strengths of deep learning models to serve as biological proxies, enabling interpretation of key gene interactions and their impact on phenotypic outcomes. The objective of this research is to refine the understanding of genetic networks in complex traits by leveraging the nuanced decision-making of advanced models.

**Results:** Testing was conducted across several computational models representing varying levels of complexity trained on gene expression datasets for the prediction of the Ki-67 biomarker, which is known for its prognostic value in breast cancer. The methodology is capable of using models as proxies to identify biologically significant genes and to infer relevant gene networks from an entirely data-driven analysis. Notably, the model-derived biomarkers (p-values of 0.013 and 0.003) outperformed the conventional Ki-67 biomarker (0.021) in terms of prognostic efficacy. Moreover, our analysis revealed high congruence between model precision and the biological relevance of the genes and gene relationships identified. Furthermore, we demonstrated that the complexity of the identified gene relationships was consistent with the decision-making intricacy of the model, with complex models capturing greater proportions of complex gene–gene interactions (61.2% and 31.1%) than simpler models (4.6%), reinforcing that the approach effectively captures biologically relevant in-model decision-making processes.

**Conclusions:** This methodology offers researchers a powerful tool to examine the decision-making processes within their genotype–phenotype mapping models. It accurately identifies critical genes and their interactions, revealing the biological rationale behind model decisions. It also enables comparisons of decision-making between different models. Furthermore, by discovering in-model critical gene networks, our approach helps bridge the gap between research and clinical applications. It facilitates the translation of complex, model-driven genetic discoveries into actionable clinical insights. This capability is pivotal for advancing personalized medicine, as it leverages the precision of deep learning models to uncover biologically relevant genes and gene networks and opens pathways for discovering new gene biomarker combinations and previously unknown gene interactions.

## Background

In the field of computational genomics, the increasing utilization of deep learning models has significantly advanced precision in linking genotypes to phenotypes [1–5]. The increasing complexity and precision of these models have ostensibly led to higher fidelity in capturing the underlying biological relationships that connect genetic inputs to observable traits [5]. This enhancement in model capability is why evaluating these models’ decision-making processes can serve as a proxy for understanding underlying biological relationships among the input features. However, these advanced deep learning models, often labeled ‘black box’ models, tend to obscure the underlying mechanisms through which their decision-making ties genetic inputs to phenotypic results. This lack of clarity impedes a thorough understanding of the intricate genotype‒phenotype relationships, which is crucial for translating these findings from research to clinical applications [6].

Our approach is designed to bridge the gap between a model’s computational predictions and the biological relevance they aim to capture, offering insights into the underlying genetic mechanics driving phenotypic variations. To achieve this goal, two fundamental steps are essential: first, identifying genes that play a crucial role in phenotypic in-model determination and, second, elucidating the complex interactions among these genes resulting in phenotypic variation.

In terms of understanding deep learning decision-making, two fundamentally different methodologies exist. The first is the post hoc approach, which encompasses various methods for interpreting a model after it has been built. This approach is more versatile and can be applied to a broader range of models [7]. The second methodology is the built-in approach, which involves integrating explainable components directly into the model’s architecture during its construction [7]. While this built-in method can provide tailored insights specific to a model architecture, it tends to be less flexible and limited in its generalizability, as it is custom designed for specific model structures [1]. Given that our approach is designed to be model-agnostic, adopting a post hoc approach is essential to ensure its broad applicability and versatility.

Previous attempts to achieve the first fundamental step of identifying model-specific phenotypically important genes have primarily utilized post hoc methodologies. Within the scope of feature importance analysis in predictive models, numerous post hoc methodologies have been developed and employed. These include SHAP (SHapley Additive exPlanations) [8], LIME (Local Interpretable Model-agnostic Explanations) [9], surrogate models [10], ALE (Accumulated Local Effects) [11, 12], and permutation-based techniques [13]. These methods have significantly enhanced our capacity to interpret model predictions by identifying key features. However, a common limitation among them is their general inability to elucidate the interactive effects of these features – specifically, how these features combine and interact to influence the model’s output.

Most approaches in computational genomics for uncovering interactions among genetic features typically involve built-in methods integrated within model architectures. These approaches include Gene‒ Gene Interaction Neural Networks (GGINNs) [14], the GenNet framework [4], Gene Network Inference (GNE) [15], Deep GONet [1], and multimodal deep learning models such as MDLCN [16]. Additionally, many of these approaches rely on preexisting gene interaction data to construct their networks, limiting their capacity to discover new phenotype-specific gene interactions or networks, namely, GenNet, GNE, MDLCN, and Deep GONet [1, 4, 15, 16]. Collectively, these methods, while insightful, are limited by their specific models or topologies, which limits their broad applicability.

This underscores the absence of a unifying post hoc methodology in computational genomics for concurrently deriving gene importance and inferring gene interactions from a model’s decision-making process with respect to a phenotype. The objective of our research was to address this gap by introducing a versatile and completely data-driven model-agnostic method applicable to any expression data, using any model, and for any binary phenotype. This method utilizes the decision-making processes of models to uncover underlying biological relationships by manipulating inputs and observing the resultant changes in outputs. This data-driven approach is compatible with any predictive model, providing a comprehensive solution to this long-standing challenge in the field.

## Materials and methods

This study draws upon two datasets from the NCBI GEO database, GSE96058 (n = 2,976) [17] and GSE81538 (n = 405) [17], encompassing gene expression data for 16,889 genes. The GSE96058 dataset was filtered to remove genes with minimal variability (standard deviation < 0.05) and those expressed at low levels (mean expression < 0.05, resulting in n = 15,132), with a focus on patients with available Ki-67 status (n = 1,363). The selected genes were subsequently identified in the GSE81538 dataset [18]. A critical aspect of both datasets is the inclusion of Ki-67 status, a marker for cell proliferation determined by immunohistochemical staining of the Ki-67 protein [17]. The Ki-67 status, which reflects the activity of the MKI67 gene, serves as an indirect indicator of the influence of gene expression on cell cycle dynamics [17, 19–21].

All code, models, and data used in our study are made publicly available on GitHub, providing transparency and reproducibility for our research. This repository can serve as a resource for other researchers and practitioners in the field, allowing them to access our methodologies, replicate our results, or even extend our work in their research endeavors. This open-source approach aligns with our commitment to collaborative and transparent scientific research, contributing to the broader scientific community’s efforts in advancing our understanding of genetic networks in cancer and other complex diseases. This research introduces a model-agnostic, data-driven methodology, structured into two phases, to bridge the gap in understanding deep learning model decision-making processes in computational genomics.

The first phase focuses on identifying genes crucial for phenotype determination using permutation testing within the context of specific models. The second phase involves constructing gene influence networks, deciphering the complex interactions among these identified genes (Figure 1). To allow for rigorous assessment of our approach, three computational models exhibiting varying levels of complexity were developed (Additional file 1, Figure S1). The aim was to delineate not only the singular impact of specific genes but also the collective influence of their interactions within a network.

**Figure 1.**
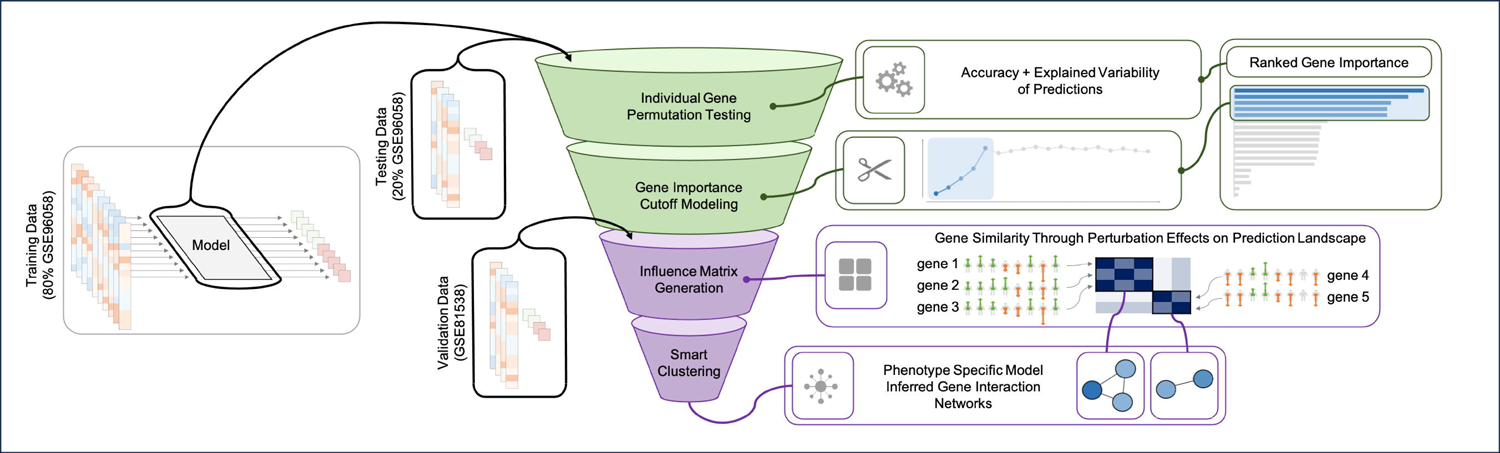
Integrative framework for model-based phenotype-specific gene analysis and influence network construction. The figure illustrates the comprehensive methodology for understanding gene contributions and interactions in phenotype determination, using a trained computational model as a proxy for biological reality. The model was trained on training data (80% of the dataset GSE96058). The approach then used this trained model, alongside testing (20% of GSE96058) and validation datasets (GSE18363), to evaluate the model’s decision-making processes. The objective is twofold: (1) to ascertain which genes are pivotal for the model to make phenotypic predictions and (2) to uncover the interactions among these important genes. This is achieved through a systematic analysis involving individual gene permutation testing and gene importance cutoff modeling, which leads to the derivation of ranked gene importance using a dual metric of accuracy and explained variance (R^2^). With the validation dataset incorporated, the methodology was extended to generate a gene influence matrix based on the similarity of average permutation effects between important genes on the predictive landscape. Smart clustering is subsequently applied to this matrix to reveal phenotype specific, model-inferred gene interaction networks, thereby illustrating the concerted gene interactions that underpin phenotypic determination as interpreted by the model.

### Model training

Three distinct models with varying degrees of complexity were trained on an identical 80% subset of the GSE96058 dataset. To manage high-dimensional data and reduce overfitting, we confined our input features to the top 100 principal components (PCs) derived from kernel principal component analysis. This dimensionality reduction was predicated on the understanding that the full complement of 15,132 genes would likely result in suboptimal classification performance for Ki-67 status within the sample size provided.

The simplest model developed was a logistic regression (LOG) model, chosen for its interpretability and baseline performance metrics. Despite its simplicity, the identification of gene importance within this model is also nontrivial due to the indirect representation of the original data by kernel-derived PCs used as input features. In contrast, the subsequent models—a multilayer perceptron (MLP) and a convolutional neural network (CNN) model—offered increased complexity and the potential for capturing more nuanced patterns within the data. Critical to our comparative analysis was the establishment of a performance benchmark to facilitate equitable evaluation across the models. This benchmark was set at an approximate 0.8 average for the area under the receiver operating characteristic curve (AUROC), with the models achieving average 5-fold cross-validation AUROC scores of 0.85, 0.85, and 0.79 for LOG, MLP, and CNN, respectively. Additionally, the models attained average area under the precision‒recall curve (AUPRC) values of 0.89, 0.89, and 0.85, respectively (Additional file 2, Figure S2). These performance indicators demonstrate the consistent predictive capability of the models and justify their use for subsequent methodological analyses.

The use of kernel PCs in conjunction with deep learning architectures was deliberately chosen to augment model performance and obscure feature importance and relationships. Despite the inherent opacity associated with such advanced models, our methodology was designed to discern gene significance and interconnectedness with respect to a given phenotype. This approach aims not only to validate the use of deep learning models as analytical proxies for genotype–phenotype relationships but also to provide a robust methodological toolkit for the broader genomic research community. By assessing the interpretability of model decisions at varying levels of complexity, we shed light on the inner workings of these models, thereby contributing to greater transparency in the field of genetic research. The performance and utility of these models, beyond their predictive accuracy, established a foundation for the next phase of our study: the comprehensive identification and analysis of biologically relevant genes and the intricate network of their interactions through permutation testing, thus setting the stage for a deeper exploration into genotype–phenotype relationships.

### Gene importance

To better understand gene significance in the context of phenotypic expression, we employed an adjusted permutation-based methodology, a technique well suited to discovering the salience of features within black box models [22]. This approach operates on the premise that the deliberate randomization of gene expression values within the dataset and the observation of how these permutations influence the model’s performance serve as indicators of gene significance. A gene whose permutation results in significant performance degradation is considered critical for phenotype prediction. Conversely, a gene whose permutation leaves model performance largely unaltered is assumed to have a minimal role in phenotypic determination. The selection of the permutation method has several advantages. Primarily, it upholds the data’s intrinsic structure, avoiding the confounding alterations that may arise from feature omission or nullification techniques [22]. This preservation is pivotal, allowing the approach to test the null hypothesis under more biologically realistic scenarios where the absence of gene expression is atypical. Additionally, permutation allows for the retention of dataset integrity, a crucial consideration when simulating the complex interplay of genetic expression.

### Individual gene permutation testing

Our permutation testing paradigm applied a dual-metric scoring system to quantify gene importance. The accuracy (*acc*) and R-squared (*R*^2^) were computed as baseline metrics from our models’ performance on the withheld test set (*acc*_*orig*_ and *R*^2^_*orig*_), which comprised 20% of the GSE96058 dataset. Subsequent permutations of each gene were conducted one hundred times (*N*), followed by a recalculation of the metrics after each permutation (*acc*_*perm, n*_ *and R*^2^_*perm, n*_). The deviations from baseline were averaged to obtain two composite scores (Δ*acc*_*g*_ and Δ*R*^2^_*g*_), which were then ranked (*rank*_*acc*, *g*_ and *rank*_*R*_2_, *g*_) then combined (*rank*_*total, g*_) and then normalized to derive a final importance score for each gene (*rank*_*g*_).

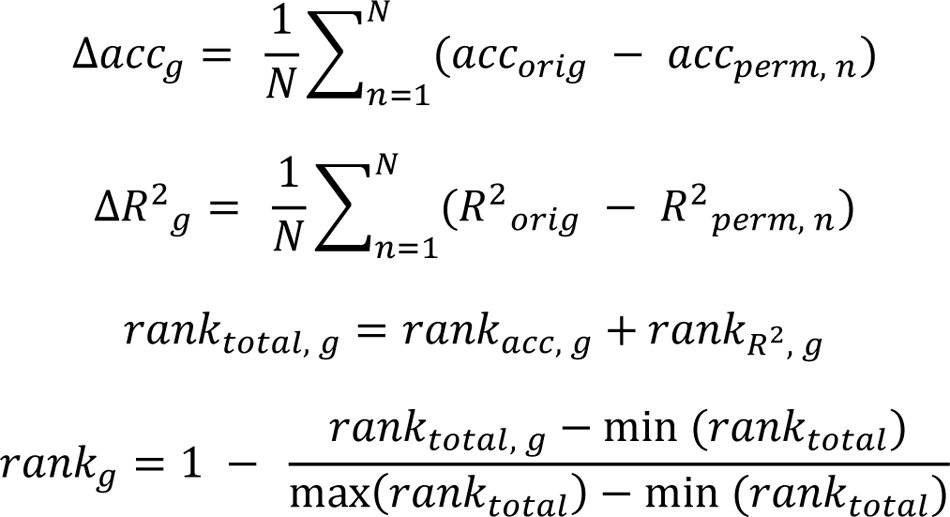

This dual-metric approach leverages the directness of accuracy and the sensitivity of R-squared to offer a comprehensive evaluation of gene importance. Although accuracy provides an immediate gauge of predictive success, it is susceptible to distortions arising from factors such as class imbalances and small individual feature effects. Conversely, R-squared provides a refined measure of the model’s predictive confidence and the subtleties of gene influence, albeit potentially overfitting to the noise within high-dimensional datasets. The concurrent application of these metrics yields a multifaceted understanding of gene importance, thus facilitating a deeper exploration into the molecular underpinnings of phenotypic manifestations.

### Gene importance cutoff modeling

To delineate a cutoff for gene importance informed by model architecture complexity, we reconstituted the architectures of the three previously developed models, this time incorporating only the most significant genes as indicated by the individual permutation testing. Notably, the input for the convolutional neural network (CNN) model was adjusted to accommodate its structural requirements through appropriate padding for nonsquare inputs. To circumvent the nondeterministic nature of deep learning models, each model underwent a series of 50 training and evaluation cycles on the training set, with successive inclusion of additional genes up to the first 100, ranked by importance. The performance of each model was quantified using the average of the area under the receiver operating characteristic curve (AUROC) and the area under the precision-recall curve (AUPRC) upon prediction on the test set (20% of GSE96058).

The rationale behind utilizing identical model architectures, sans PCA, was to permit the complexity inherent in these models to influence the determination of this gene importance threshold. This approach represents a departure from traditional methodologies that typically employ simpler models and overlook the effect of model architecture on the significance cutoff. By analyzing the progression of AUROC scores corresponding to the incremental addition of genes, we identified the cutoff as the point where further inclusion of genes did not appreciably enhance the model’s performance, operationalized as the first instance when the AUROC reached within 0.025 of the highest value observed across the set of 100 top genes used. This process enabled us to pinpoint the optimal number of genes that strike a balance between contributing to phenotype prediction and the complexity introduced by the model architecture. By adopting this strategy, we intended to provide a cutoff that is not only reflective of gene significance but also cognizant of the computational framework within which the analysis is conducted.

### Assessment of critical gene selection

Then, to evaluate the efficacy of gene selection, a dual approach was used. First, we compared the modeling performance of the top critical genes identified through various feature selection methodologies, including surrogate modeling, SHAP built on a surrogate model, SHAP with inverse mapping from PC space to the original feature space, LIME, and ALE-based feature selection (Additional file 6, Table S1). These feature selection methodologies were run on the LOG, MLP, and CNN models, and their top critical gene selections were tested over 100 simulations. Their AUROC scores were compared to the AUROC generated by 100 simulations of the top critical genes selected by our individual gene permutation approach. This evaluation focused on both the average predictive performance and the stability of these scores across simulations, which are key factors for robust model deployment and feature selection methodology.

The second aspect of our evaluation focused on the biological relevance of the critical genes selected by our approach. Initially, we conducted a thorough literature review of each critical gene identified by modeling approaches (LOG, MLP, and CNN) to determine their association with breast cancer (BC), their known or proposed role as prognostic biomarkers (BM) for breast cancer, and their use as therapeutic targets (TT) for breast cancer. This literature review was conducted with a focus on breast cancer, as the Ki-67 status is used as a biomarker to determine prognosis and treatment, and both datasets included breast cancer patients [17]. In addition to the literature review, we generated a custom biomarker to further assess the biological significance of these genes. The biomarker was calculated based on the significant differential expression of genes in the context of Ki-67 status. For each gene, if its expression was significantly different between two groups (Ki-67 positive and negative), it was either added to or subtracted from the biomarker value, depending on whether it was overexpressed or underexpressed in the group with high Ki-67 status. This method allowed us to create a biomarker reflecting the aggregate influence of multiple significant genes.

This custom biomarker was used to perform a survival analysis on the full GSE96058 dataset. The effectiveness of our biomarkers, generated from critical values for each model architecture, was measured by their ability to distinguish between two survival groups using the log-rank test p-value for comparison. This performance was benchmarked against that of the widely recognized Ki-67 status biomarker [17], which our models aimed to predict. This comprehensive methodology, combining predictive modeling performance with biological relevance analysis, provided a robust evaluation of the critical gene selection in our study with respect to both other feature selection methodologies and ensured the biological relevance of our approach.

### Influence networks

We propose that influence networks are crucial for delineating the intricate connections between genes and their collective influence on phenotypic traits. These networks are constructed by analyzing gene permutations within a model-specific testing framework, focusing on the effects of individual gene expression changes on model predictions. This involves adjusting the expression of a single gene and observing the resulting shifts in the predictive model across different patient samples. The aim is to uncover genes with similar or opposing impacts on predictions, hinting at potential biological ties to the studied phenotype. This method delves deeper than just assessing individual gene importance; it explores the underlying genetic interdependencies that drive complex phenotypic expressions.

Our influence networks, generated by a completely data-driven and model-centric approach, avoid preexisting biological hypotheses, allowing for novel gene network discovery relevant to the specific phenotype of interest. This flexibility enables the discovery of both established and new gene interactions directly from model behavior and patient data, a significant advantage when investigating less understood genetic interactions. To construct these influence networks, two fundamental steps are necessary: first, generating influence matrices to illustrate the pairwise relationships of effects of gene permutations on model predictions, and second, forming clusters of genes based on their influence, indicating that genes influencing predictions in similar or opposite ways may be related to similar biological processes.

### Permutation-based prediction effect similarity (influence matrix)

To discern gene similarity in terms of their impact on model predictions, we conducted a systematic analysis to gauge the average change in prediction probability for each of the top 100 genes. This extensive approach, focusing on a wider gene spectrum instead of solely the previously identified critical genes, was aimed at capturing a more comprehensive perspective of potential genetic interactions. The rationale for examining the top 100 genes, beyond the critical gene set, stemmed from multiple considerations. Primarily, critical gene selection, though invaluable, originated from models excluding the kernel PC step, differing from the original trained models that embody the full complexity of phenotype prediction. Moreover, examining a uniform number of genes across models facilitates the comparability of influence networks derived from diverse modeling methodologies.

Leveraging the original, trained models, we implemented permutation testing on the test set. Each of the top 100 genes, ranked by importance, was individually permuted in 100 iterations. Following kernel PC transformation, the test set was subjected to phenotype prediction using the model, which enabled us to capture the average deviation in each patient’s predicted phenotype due to gene permutation. This process involved two sequential steps:

1. **Computation of average prediction change:** For each gene *g* within the data frame of all genes *X* that are also in the top 100 important genes, we permute its normal expression profile while keeping all other genes the same (*X*_*perm, g*_) and observe the resultant variation in phenotypic predictions (Δ*P*_*g*_) for all patients using model *M* and the kernel PC reduction (*PCA*). This process was repeated over 100 simulations *S* to ensure robustness in the assessment of the influence of each gene. The rationale for using the original PCA-transformed data was to maintain fidelity to the model interpretive framework, thereby ensuring that the influence matrix accurately reflected the learned associations of the model.

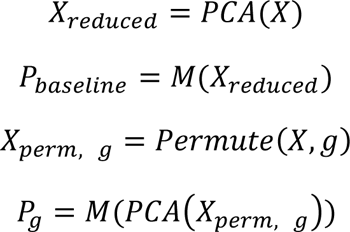

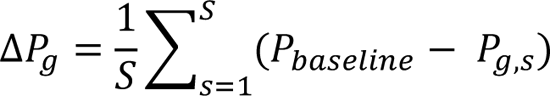
2. **Pairwise gene correlation analysis:** Upon establishing the average prediction changes, we computed pairwise correlations between the genes. This analysis aimed to reveal genes whose permutations had similar or diametric effects on the phenotype, suggesting a potential interactive relationship within the predictive model. Specifically, for genes *g*_1_ and *g*_2_, the correlation of their respective Δ*P* vectors was computed to evaluate their interplay.

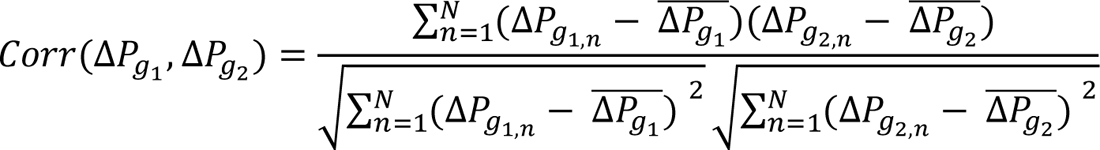

Here, *N* represents the number of patients, and ̅*P_g_* denotes the mean of the prediction vector change for gene *g*. The final influence matrix is a pairwise matrix of all combinations of gene pairs (within the top 100 genes) generated by each model.

The influence matrix, derived from the average prediction changes and subsequent correlation analysis, serves as a quantitative scaffold for modeling gene interactions. It enables a data-driven exploration of genetic interplay, potentially uncovering both known and novel genetic associations. By adopting this thorough empirical approach, we offer a novel lens through which researchers can explore model-inferred gene interactions. This not only aids in validating the biological relevance of the predictive models but also paves the way for discovering new genetic connections, which are crucial to understanding complex phenotypes. The influence networks constructed through this methodology thus represent a significant stride toward aligning computational predictions with biological functionality.

### Smart clustering of influence matrices

Following the derivation of the influence matrices, the next objective was to cluster genes to elucidate their collective impact on phenotypic traits. This was aimed at partitioning the top 100 gene sets into clusters (based on the influence matrix) that would represent their mutual interactions as determined by the predictive model.

To achieve this, a process to identify the optimal clustering strategy was developed. The range of clustering algorithms tested included k-means, agglomerative clustering, spectral clustering, Gaussian mixture models (GMMs), affinity propagation, and OPTICS (ordering points to identify the clustering structure). Recognizing the complexity of the gene interaction data, we first transformed the influence matrix to absolute values. This preprocessing step was performed to ensure that the influence of genes with diametric effects on the phenotype was accurately captured, acknowledging the dual nature of genetic regulation.

Each clustering algorithm underwent rigorous parameter optimization. A large set of parameter configurations for each clustering methodology was searched (Additional file 7, Table S2). This was particularly crucial for algorithms prone to variability in cluster assignments. To establish the most representative clustering outcomes, we ran multiple iterations (50 per parameter set) to assess the stability and reliability of the results. The primary metric for evaluating clustering quality was the Dunn index, which was chosen for its ability to identify well-separated and compact clusters [23, 24]. This metric aligns well with the inherent properties of genetic data, where biological processes often manifest as tightly knit gene clusters. Although the Dunn index was the preferred metric, the flexible approach also accommodated alternative metrics such as the silhouette score, Davies–Bouldin score, and Calinski–Harabasz score, enabling adaptability to different analytical requirements. Once the optimal parameters were determined for each clustering method, the most effective algorithm was subjected to additional validation through 1000 simulations under the best parameter setup. This step accounts for stochastic elements in some algorithms, aiming to select a clustering outcome that consistently achieves the highest Dunn index or the best score according to the chosen metric.

This ‘smart clustering’ strategy was used because different models may reveal distinct patterns of gene interactions. By evaluating multiple clustering methodologies and parameters, we ensured the selection of the most fitting clustering arrangement for the specific data, phenotype, and model under investigation. This rigorous process not only enhances the precision of the resulting gene influence networks but also promotes accessibility and reproducibility for researchers, fostering further biological discoveries within a data-driven paradigm.

The transformation of gene clusters into influence networks involved representing each gene as a node in a graph, with edges symbolizing the interactions between genes as inferred from the influence matrix. To impartially represent both synergistic and antagonistic gene relationships, the absolute values from the influence matrix were used for edge construction. This method allowed us to depict genes with opposing effects on the phenotype as interconnected within the network. The size of each node corresponded to the gene’s relative importance, with edge colors—black for synergistic interactions and red for antagonistic interactions—indicating the nature of the gene relationships. Furthermore, nodes were color-coded (blue for critical genes and green for noncritical genes within the top 100 genes) to distinguish their categorization. For enhanced clarity and interpretability of the network, each cluster was subjected to selective pruning of edges. Edges were sorted by their weight and incrementally removed as the process continued until it disrupted the network’s connectivity. Notably, if a cluster had a small number of nodes (less than 10), the pruning was adjusted to retain at least two edges per node. This modification was implemented solely for the ease of visualizing these networks, ensuring that even smaller clusters remained interpretable. It is important to note that while this visualization technique displays the tightest connections within each cluster, in reality, each cluster is fully connected. The visual representation aims to emphasize the most significant interactions within these networks.

### Individual cluster analysis

The next step was to conduct an examination of the individual clusters formed in the influence networks to determine the biological relevance and structural integrity of these clusters. By employing a two-pronged approach, we aimed to validate the computational findings against established biological databases and frameworks. This step was essential for determining whether the gene clusters identified by the model mirrored natural gene structures and were functionally relevant to the phenotype being studied.

1. **Overlap with GeneMania clusters:** This research was extended to compare the gene clusters identified through the model with those present in GeneMania, a comprehensive database for functional interaction networks among genes [25]. This comparison was pivotal in determining whether our model-derived clusters have biological parallels in existing gene interaction databases. A high degree of overlap would indicate that the computational clusters are not only data-driven but also biologically relevant, reflecting actual gene interactions found in nature.
2. **Gene Ontology (GO) enrichment analysis:** An enrichment analysis of Gene Ontology (GO) terms within the clusters was conducted. This analysis focused on identifying overrepresented biological processes, cellular components, or molecular functions in each cluster [26]. The enrichment of specific GO terms in a cluster suggested that the genes grouped together in our influence networks were functionally coherent and relevant to the phenotype of interest.

Together, these analyses provide dual validation of the data-driven gene clusters. By comparing the clusters with established gene interaction networks and biological functions, we demonstrated that the networks identified by our models are not only computationally robust but also fundamentally aligned with natural biological systems and relevant to the phenotype under study. This multifaceted approach underscores the biological significance of the computational groupings derived from the influence networks.

### Combined cluster analysis

Finally, the focus shifted toward exploring the variety and types of gene relationships that are discovered by our influence networks, which were generated by various modeling architectures. This exploration aimed to identify and understand the types of relationships each model architecture could reveal. The process entailed the following steps for the influence clusters identified by each model:

1. **GeneMania comparison:** GeneMania was utilized to examine the types of gene‒gene relationships present within each cluster identified within the model. This analysis helped in understanding the nature and dynamics of gene interactions as revealed by the influence networks derived from different modeling architectures. This comprehensive database includes information on various types of gene‒gene relationships, such as coexpression, genetic interactions, physical interactions, colocalization, pathway participation, predicted interactions, and shared protein domains [25]. For the GeneMania comparisons, the search parameters used were specifically configured to input the genes from each cluster, with the maximum number of resulting genes set to zero. This restriction ensures that only direct (1-hop neighbor) relationships are analyzed for simplicity. Additionally, the maximum number of resultant attributes was capped at 10 (default), and the weighting scheme was set to be based on the query genes, thus maintaining a focused and relevant output.
2. **Relationship aggregation:** We aggregated the types of relationships for the clusters identified by each individual modeling methodology. This aggregation provided a summary of interaction types prevalent within the clusters of a given model, offering insights into the nature of the relationships inferred by that particular model.
3. **Intermodel comparison:** A comparison among these aggregated interaction profiles was performed across models. This comparison was aimed at discovering how different modeling architectures might vary in uncovering the complex web of gene interactions and their potential implications for understanding the phenotype of interest.

Through this analytical approach, we sought to delve more deeply into the kinds of gene relationships that our influence networks, borne out of diverse modeling architectures, could elucidate. This not only offered a nuanced understanding of model-specific capabilities but also enriched our comprehension of the biological networks underlying the studied phenotypes.

## Results

To assess the applicability of the models in the context of breast cancer prognosis, their performance in predicting Ki-67 status, a critical biomarker in breast cancer, was evaluated [19]. The trained models (LOG, MLP, and CNN) were tested on two datasets: 20% of the GSE96058 dataset was used as an internal test set, and GSE81538 was used as an external validation set. The AUROC values were 0.78/0.80 (LOG), 0.79/0.82 (MLP), and 0.77/0.73 (CNN) respectively (Additional file 3, Figure S3). While the CNN model showed a decline in performance on the external dataset, it remained well above the baseline AUROC of 0.50, indicating its predictive validity, albeit less effective than the LOG and MLP models. These models, which are designed with varying complexities, were tasked with predicting Ki-67 status, a well-known biomarker in cancer. The objective was to show that it is possible to harness the interpretative power of these complex models to unravel the intricate features essential for cancer prognosis. The aim was to uncover not only key prognostic factors but also the complex gene interactions that underpin cancer progression. This approach was intended to provide a deeper understanding of the biological underpinnings that drive cancer behavior, leveraging the models’ analytical capabilities to illuminate the underlying genetic networks.

### Assessment of critical gene selection

The results of the methodology indicated that 44, 21, and 49 of the top genes were needed for the LOG, MLP, and CNN models, respectively, for plateauing modeling performance (Additional file 4, Figure S4). This implementation provided insights into the optimal number of genes required for effective model performance. We substantiated our critical gene selection approach on two fronts: first, by demonstrating that the genes identified as critical through our modeling techniques effectively elucidate Ki-67 status, surpassing previous methodologies; and second, by confirming the biological significance of these genes in the context of breast cancer progression and survival.

1. **Model-based justification:** The comparative analysis of feature selection methodologies validated the efficacy of our full permutation feature selection approach (Figure 2). Compared with established methods such as SHAP inverse mapping, ALE, surrogate model, SHAP with surrogate model, LIME-based approaches, our method demonstrated either greater or marginally greater performance in predicting Ki-67 status. It was tested across all three models (LOG, MLP, and CNN) using the critical gene count determined by our cutoff, and our approach yielded the highest average AUROC score of 0.823 and one of the lowest standard deviations in these scores (0.015), indicating the method capability and stability in feature selection for model predictions (Figure 2).
2. **Biological relevance:** The evaluation of the LOG, MLP, and CNN models yielded significant insights into the biological relevance of their critical genes in the context of breast cancer (Figure 3). Through this method, we discovered that a considerable percentage of the genes (84% for LOG, 80% for MLP, and 67% for CNN) are known to be associated with breast cancer, as verified by an extensive literature review (Additional files 8-10; Tables S3-S5). Furthermore, our findings suggest that 61% of the genes identified through both the LOG and MLP models and 53% identified through the CNN model are recognized or proposed prognostic biomarkers for breast cancer. The analysis also revealed that 36%, 53%, and 35% of the critical genes identified by the LOG, MLP, and CNN models, respectively, are recognized or proposed as therapeutic targets in breast cancer treatment (Figure 3).

**Figure 2.**
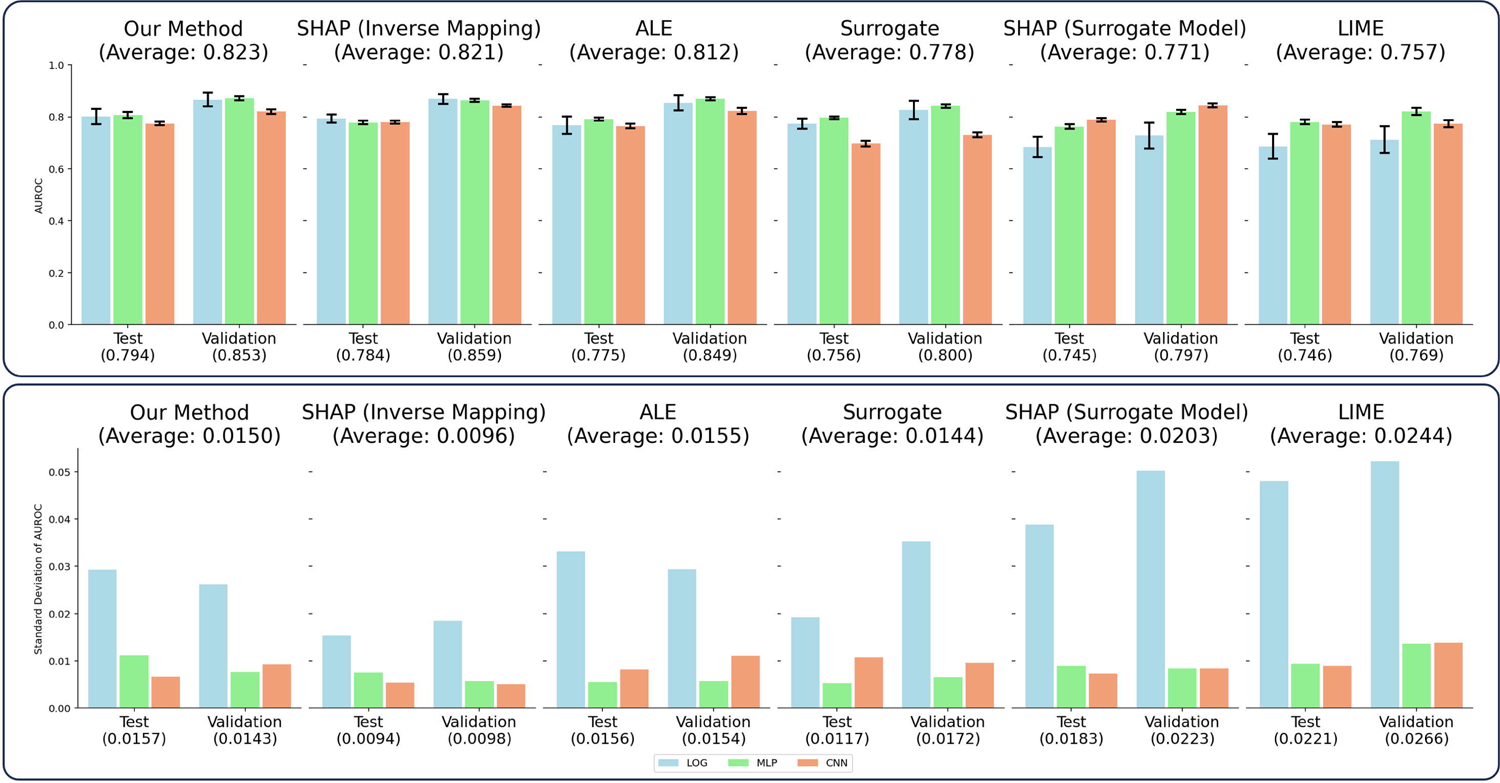
Comparative performance of feature selection methodologies on fixed-size feature sets across model architectures. This figure illustrates the performance impact of distinct feature selection methodologies on the three deep learning architectures when constrained to the critical number of top features (LOG – 44, MLP – 21, CNN – 49). The feature sets, while equal in number for each model type, are composed of different genes, and each set is selected based on the rankings provided by the respective feature selection method. AUROC performance (top panel): The graph shows the average AUROC scores for each methodology, representing the discriminative power in predicting Ki-67 status. These scores are averaged across 100 simulation runs to ensure robustness in the performance evaluation. AUROC stability (bottom panel): The corresponding standard deviations of the AUROC scores, showing the stability and repeatability of each feature selection method. The results underscore the efficacy of our method, which not only achieves superior average AUROC scores but also demonstrates one of the lowest standard deviations, confirming its robustness and consistency in identifying the most predictive features for Ki-67 status.

**Figure 3.**
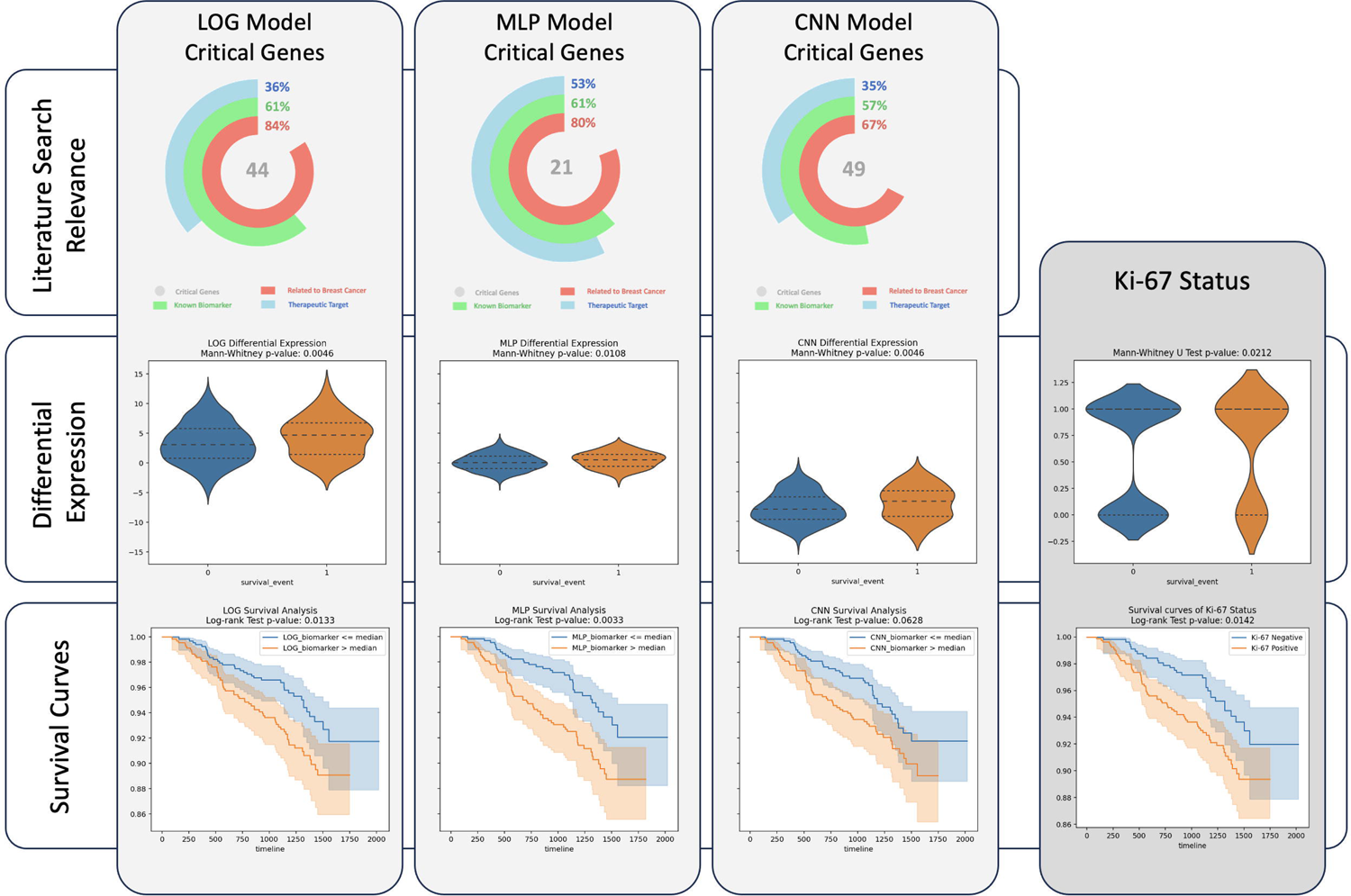
Biological relevance of critical genes identified across model architectures. This figure presents a multifaceted evaluation of the biological significance of critical genes identified by our feature selection methodology, as applied to logistic regression (LOG), multilayer perceptron (MLP), and convolutional neural network (CNN) models. Literature search relevance (top panel): The donut charts illustrate the literature search results for the critical genes, with the central gray number indicating the total count of critical genes identified. The outer colored rings represent the percentage of genes associated with breast cancer (red), known or proposed as prognostic biomarkers (green), and known or identified as potential therapeutic targets (blue). Differential expression analysis (middle panel): Violin plots displaying the differential expression of a custom biomarker panel derived from critical genes with significant differential expression according to Ki-67 status. The plots contrast survival groups with the Mann‒Whitney P value indicating expression differences between the two groups. Survival analysis (bottom panel): The survival curves generated from model-specific biomarkers were used to assess the prognostic value of the identified critical genes. The curve separation and the log-rank test p-value underscore the significance of the biomarkers in survival outcomes. Baseline (far right column): This column shifts the focus to the established Ki-67 biomarker. The differential expression by survival groups and their corresponding survival curves are depicted, serving as a reference point for the critical gene biomarker panels derived from our evaluated models.

Notable among the therapeutic target genes associated with breast cancer identified through this approach with the LOG model are *TFPI2* [27–29], *TROAP* [30–33], and *TONSL* [34–36]. In the MLP model, critical genes such as *TNNT1* [37, 38] and *AQP7* [39–41] were also identified as potential therapeutic targets. Five genes (*TACC3* [42–45], *CDC20* [46–50], *FOXI1* [51–53], *KIF11* [54–57], and *MKI67* [17, 19–21]) were identified as critical in both the LOG and MLP models and have been proposed to be therapeutic targets, with *MKI67* being particularly noteworthy because it encodes the Ki-67 protein. Additionally, our analysis employing the CNN model identified several key genes as potential therapeutic targets for breast cancer. These genes include *GRIK3* [58, 59], *BEX2* [60–64], *AGTR1* [65–68], and *PAX2* [69–71]. Significantly, many of the genes underscored in the literature review as therapeutic targets, including TACC3, MKI67, TFPI2, TNNT1, FOXI1, KIF11, GRIK3, BEX2, and AGTR1, are featured prominently in the top 10 critical genes across the three model (Table 1). A comprehensive overview of all critical genes identified through our methodologies utilizing the LOG (Additional file 8, Table S3), MLP (Additional file 9, Table S4), and CNN models (Additional file 10, Table S5) and their respective relationships with breast cancer — whether as established relationships, proposed or recognized prognostic markers, or as potential therapeutic targets — is included.

**Table 1.**
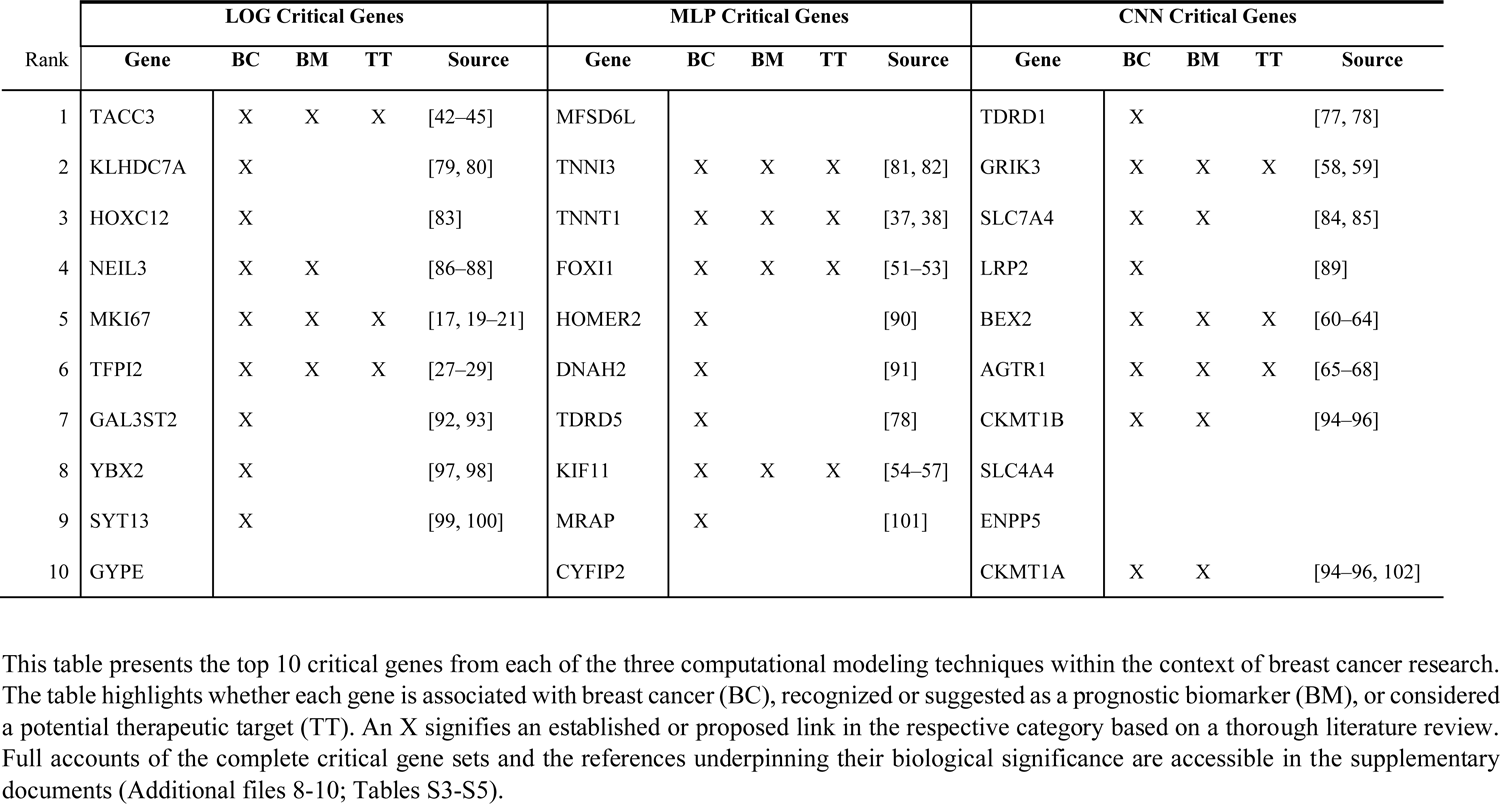
Summary of literature search of the top 10 critical genes identified by evaluation of each model.

Furthermore, to demonstrate the biological relevance of the identified critical genes beyond the support of the literature and from a prognostic standpoint, we utilized the GSE96058 dataset, which includes survival data, to develop custom biomarkers. These biomarkers, derived from the critical genes identified by our top two models (LOG and MLP), markedly outperformed the Ki-67 status as a biomarker, a widely recognized and utilized indicator of breast cancer prognosis. Survival analysis revealed significant differences (p-value < 0.05 in the log-rank test) for biomarkers generated by the LOG (0.0133) and MLP (0.0033) models, both surpassing the performance of the Ki-67 biomarker (0.0142), while the biomarker from the CNN (0.0628) model exhibited a near-significant result (Figure 3).

This evidence underscores the effectiveness of our approach in using models as proxies to elucidate their biological underpinnings, particularly in the context of breast cancer prognosis. This highlights how our methodology not only identifies key biomarkers but also demonstrates the intrinsic relationship between the performance of these models and their fidelity in capturing the nuances of underlying biological systems. The higher the model’s performance is, the more accurately it reflects complex biological realities, which in turn enhances our ability to extract meaningful biological insights from these models. This interplay between model performance and biological discovery validates the significant role of our approach in advancing our understanding of breast cancer from a data-driven and biologically informed perspective.

### Individual cluster analysis

The next phase involved developing influence matrices for the top 100 genes from each model architecture (LOG, MLP, and CNN), revealing the gene interactions within our predictive models. These matrices encapsulate the weighted mean influence of gene permutations, averaged across the testing (273 patients) and validation (405 patients) sets. This balanced approach accounts for variances in sample sizes between datasets. Utilizing our ‘smart clustering’ algorithm, we delineated clusters within these matrices, optimizing classification to accurately represent the genetic interplay learned by our models. These clusters illustrate our understanding of how groups of genes collectively influence predictive outcomes, providing a graphical exploration of their synergy or antagonism (Additional file 5, Figure S5).

**1. Overlap with GeneMania clusters:** A critical part of our analysis involved the comparison of three distinct clusters (one from each model) with the gene interaction networks from GeneMania (Figure 4). This cross-referencing revealed impressive concordance; every gene within our clusters was also represented in GeneMania’s networks, thereby substantiating the biological relevance of our model-derived clusters. Particularly notable is the alignment observed in the LOG and MLP model-derived clusters with those in GeneMania. For instance, genes such as *CIT, TRAIP, TONSL, NEIL3, TIMELESS, SKA1*, and *RECQL4* in the LOG model and *MND1, POLQ, CDCA2, ESPL1*, and *PKYT1* in the MLP model showed consistent peripheral positioning in both our influence networks and those established in GeneMania. The specific CNN model-generated influence network, although smaller, also displayed significant concordance, with genes such as *FOS, FOSB, IEE2, JUNB, ERG1*, and *ERG3* mirroring the known GeneMania network (Figure 4). This correlation between our model-generated networks and established genetic interactions attests to the precision and biological relevance of our method.
**2. Gene Ontology (GO) enrichment analysis:** This analysis of clusters from the LOG and MLP models revealed a significant enrichment of processes intimately linked to cancer survival and Ki-67 status (Table 2). GO terms such as “mitotic nuclear division,” “chromosome segregation,” and “sister chromatid segregation,” known for their roles in cell proliferation and genomic stability [72, 73], were notably prevalent. Additionally, terms related to the immune response, such as “response to type I interferon” and “regulation of viral process”, which are known to impact cancer progression [74, 75], were also enriched. Interestingly, a cluster from the CNN model evaluation exhibited enrichment for “recombinatorial repair” (FDR: 5.73e-2), just beyond the significance threshold, further indicating its relevance in cancer progression [76]. This congruence between the identified GO terms and recognized cancer-related biological processes demonstrated that our computational approach effectively leverages model decision-making to decode complex biological interactions related to Ki-67 status. The top 10 GO terms for three clusters exhibited significant enrichment (Table 2). The clusters derived from our predictive models not only demonstrated alignment with known biological networks but also resonated with key functional pathways central to cancer survival and Ki-67 status (Table 2).

**Figure 4.**
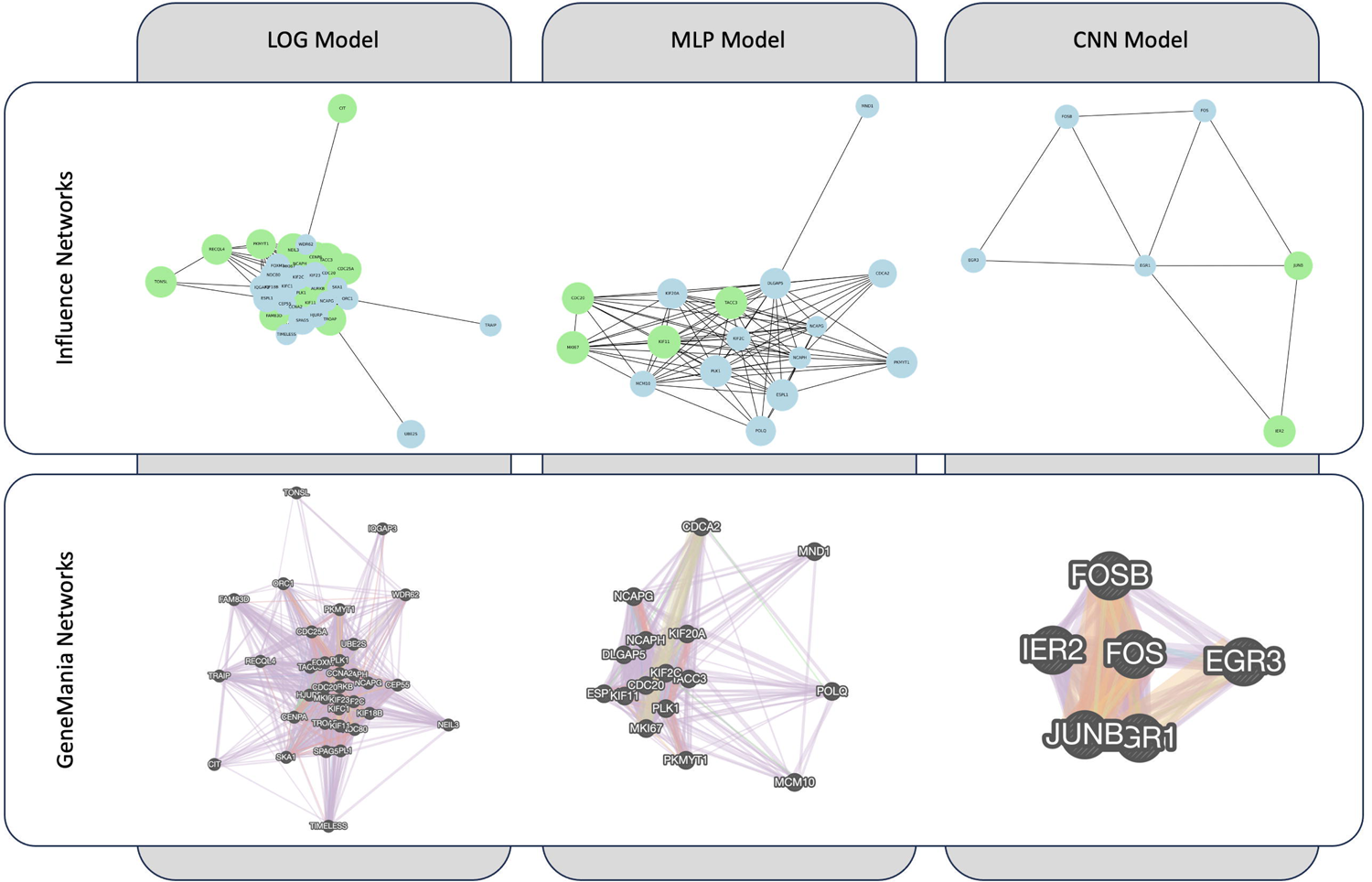
Comparative visualization of example model-inferred gene influence networks and corresponding biological networks. Generated influence networks (top panel): The diagrams illustrate an example of influence networks as inferred by three different computational models. Nodes represent genes, with proximity indicating stronger influence relationships—denoted by black edges for positive correlations (red for negative). Node color indicates gene ranking, with light blue for the top 100 genes and green for those identified as critical. Node size is proportional to gene importance within the model. GeneMania networks (bottom panel): Corresponding GeneMania networks for the genes in the specific influence network identified above. These networks show various types of gene–gene interactions, including coexpression (purple), physical interactions (red), genetic interactions (green), predicted relationships (orange), colocalization (blue), pathway interactions (cyan), and shared protein domains (tan). These networks serve as a biological validation and comparison layer for the computational influence networks shown above, offering a holistic view of gene interrelationships in a biological context. Additional file 11, Table S6 provides an extended review of all networks identified within the top 100 genes for each model evaluation and their GeneMania counterparts.

**Table 2.** Statistically significant Gene Ontology (GO) terms derived from the discovered influential gene networks.

| LOG (Cluster 1) |  |  | MLP (Cluster 5) |  |  | MLP (Cluster 7) |  |  |
| --- | --- | --- | --- | --- | --- | --- | --- | --- |
| Function | FDR | Coverage | Function | FDR | Coverage | Function | FDR | Coverage |
| mitotic nuclear division | 1.19e-20 | 18/270 | mitotic nuclear division | 2.11e-13 | 11/270 | response to type I interferon | 2.01e-19 | 10/88 |
| chromosome segregation | 7.88e-18 | 16/248 | mitotic sister chromatid segregation | 4.77e-10 | 8/137 | cellular response to type I interferon | 2.01e-19 | 10/88 |
| sister chromatid segregation | 1.24e-16 | 14/176 | chromosome segregation | 4.77e-10 | 9/248 | response to virus | 5.68e-16 | 10/197 |
| nuclear chromosome segregation | 1.24e-16 | 15/236 | sister chromatid segregation | 2.06e-9 | 8/176 | response to viral genome replication | 7.17e-11 | 7/94 |
| mitotic sister chromatid segregation | 2.24e-14 | 12/137 | nuclear chromosome segregation | 1.76e-8 | 8/236 | negative regulation of viral process | 8.61e-11 | 7/102 |
| spindle | 1.42e-11 | 12/236 | regulation of nuclear division | 5.29e-7 | 6/106 | regulation of viral process | 8.61e-11 | 8/210 |
| microtubule cytoskeleton organization involved in mitosis | 6.24e-11 | 10/133 | metaphase/anaphase transition of cell cycle | 1.20e-6 | 5/53 | regulation of symbiotic process | 8.71e-11 | 8/221 |
| spindle organization | 3.41e-8 | 9/172 | mitotic sister chromatid separation | 1.20e-6 | 5/54 | viral genome replication | 8.71e-11 | 7/106 |
| condensed chromosome | 4.43e-8 | 8/114 | regulation of mitotic sister chromatid separation | 1.20e-6 | 5/57 | regulation of viral life cycle | 8.64e-10 | 7/149 |
| chromosomal region | 4.57e-8 | 10/266 | regulation of chromosome separation | 1.42e-6 | 5/59 | viral life cycle | 4.82e-8 | 7/267 |

This table presents the top 10 GO terms from three distinct gene clusters. These terms are associated with essential biological functions in cell division and the immune response, which are critical for understanding Ki-67 status in cancer prognosis. The false discovery rate (FDR) values show the reliability of the associations, and the coverage indicates the number of genes within each cluster that are linked to the respective GO term. Clusters not shown lacked significant GO terms (FDR < 0.05) or associated biological processes according to GeneMania data.

### Combined cluster analysis

In the analysis of combined cluster gene interactions, we aimed to evaluate the nature of relationships within clustered gene networks identified via our models compared to established biological interactions in GeneMania. Our methodological approach enabled us to discern distinct patterns in the types of gene‒gene relationships reflected by the different models. The LOG model primarily captured simpler gene–gene relationships, such as coexpression and colocalization. Only an average of 4.6% of the interactions across clusters represented more complex interactions, such as physical interactions, genetic interactions, predicted associations, pathway involvements, or shared protein domains (Figure 5).

**Figure 5.**
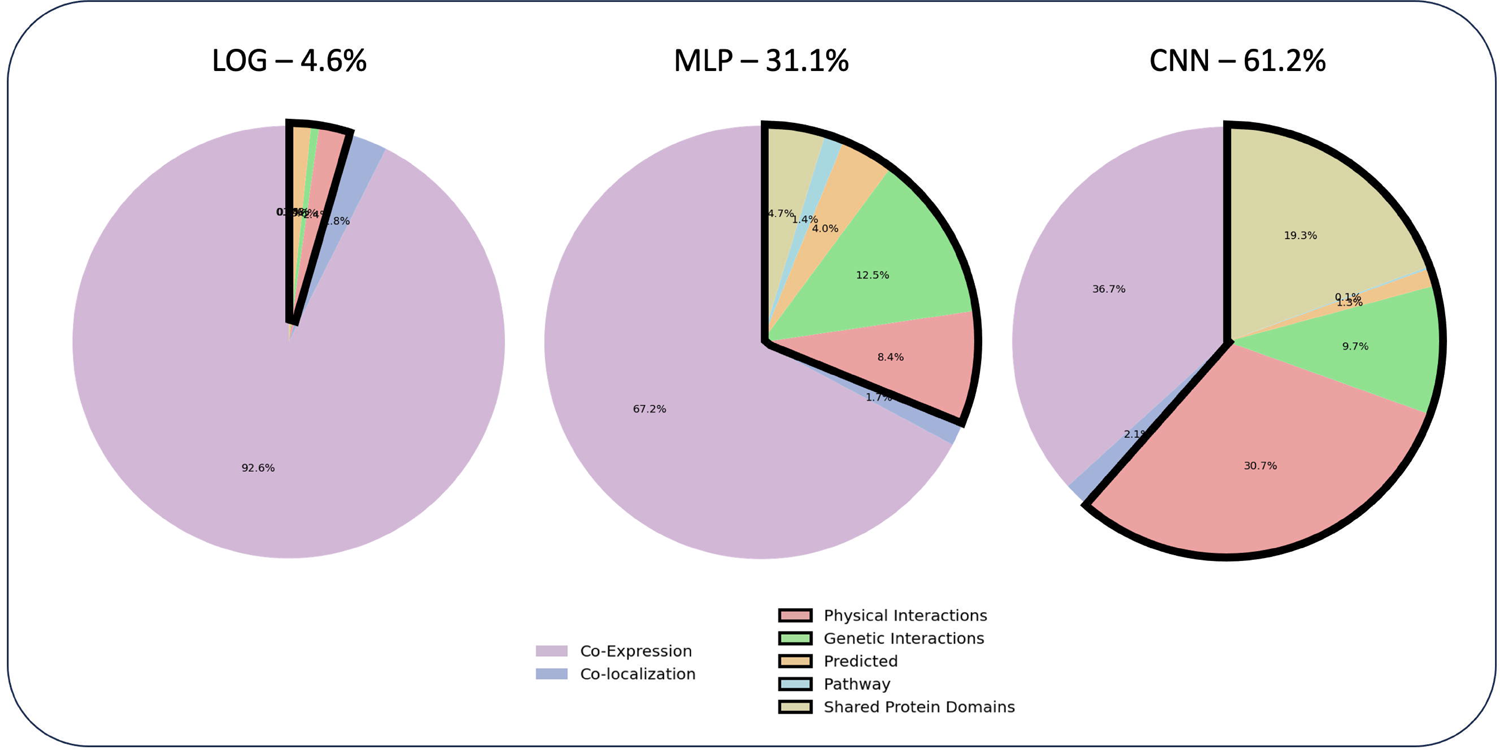
Distribution of interaction types averaged across gene clusters by model. The figures above display the proportional breakdown of gene‒gene interaction types within the clusters generated by the three different models as given by GeneMania searches. The LOG model (left) primarily captures coexpression (purple) and colocalization (blue) interactions, with a minor representation of complex interaction types (4.6%). In contrast, the MLP (middle) and CNN (right) models show an increased proportion of complex interaction types, including physical interactions (red), genetic interactions (green), predicted interactions (orange), pathway interactions (cyan), and shared protein domains (brown), making up 31.1% and 61.2% of the average across all of their respective clusters. This suggests a relationship between the complexity of the model and the sophistication of the biological relationships our approach can infer.

In contrast, the MLP and CNN models, which are better suited for capturing intricate relationships, showed a marked increase in the complexity of identified interactions. The MLP models exhibited an average of 31.1% of such complex relationships, while the CNN models demonstrated an even greater proportion, with an average of 61.2% (Figure 5).

These findings illustrate that as the complexity of computational models increases, their ability to reflect the nuanced interplay of gene interactions observed in biological systems correspondingly increases. The intricate relationships discerned by the more sophisticated MLP and CNN models are in line with expectations, suggesting that these models are more adept at unraveling the elaborate web of gene interactions that occur in nature.

## Discussion

Our research presents a novel computational methodology that considerably enhances our understanding of gene interactions, especially in the context of complex diseases such as cancer. This model-agnostic approach breaks new ground by transcending the traditional confines of predictive modeling, offering universal applicability across various computational frameworks, from logistic regression to the complexities of deep neural networks. A key strength of this approach lies in its ability to reveal the inner workings of complex models by methodically controlling inputs and observing changes in predictions. This aspect is crucial because it means that the specifics of the model’s internal architecture are irrelevant to the methodology’s effectiveness. It can be applied to any model designed for predicting binary outcomes, thereby offering a wide range of applicability. This input‒output-based approach is particularly powerful in unmasking the nuanced decision-making processes of these models, allowing for a deep dive into the relationships and interactions within genetic networks.

The robustness of this methodology is evidenced by its ability to extract biologically meaningful relationships from the models used. These models not only demonstrate predictive accuracy but also reflect fidelity to complex biological interactions. The enrichment of cancer survival- and Ki-67-related GO terms within their clusters and the identification of clinically relevant biomarkers and therapeutic targets such as MKI67 validate the alignment of this approach with biological realities. Moreover, the analysis shows that as models increase in complexity, such as MLPs and CNNs, they are better equipped to capture a wider spectrum of genetic interactions. This finding suggests that the sophistication of a model is proportional to the depth of biological insights it can provide. As such, this methodology is well suited to evolve with the advancing landscape of computational biology, promising richer insights as models become more complex. While the computational demands of our approach are significant, its effectiveness more than compensates for these requirements. In resource-limited settings, a hybrid approach incorporating SHAP interaction-based feature importance for initial feature determination, followed by our gene influence network analysis, is recommended. This strategy maintains a high level of biological insight extraction while managing computational constraints. In its current design, our approach is limited to models based on predictions of binary outcomes. While this allows for broad applicability in many biomedical scenarios, future expansions to accommodate multiclass and continuous outcomes could further enhance its utility and scope in genomics research.

## Conclusions

In conclusion, our study not only demonstrates the efficacy of using advanced computational models to understand complex genetic networks but also opens new avenues for more interpretable and biologically relevant applications of these models in genomics. By leveraging model decision-making processes, this methodology identifies critical genes and interactions, offering valuable insights into the biological rationale behind these decisions. This capability is pivotal in advancing personalized medicine, as it leverages the precision of deep learning models to uncover biologically relevant genes and gene networks, fostering the discovery of new biomarker combinations and previously unknown gene interactions.

## Declarations

### Ethics approval and consent to participate

Not applicable.

### Consent for publication

Not applicable.

### Availability of data and materials

This study analyzed publicly available, de-identified human gene expression datasets from the NCBI GEO database (accession numbers GSE96058 and GSE81538). The datasets generated and/or analysed during the current study are available in the Gene_Network_Project repository, https://github.com/ok-tsar/Gene_Network_Project.

## Authors’ contributions

RS and XC designed the project. RS and XC collected the data. RS performed the experiments and analyzed the data. XC and ZZ supervised the project. RS drafted the manuscript. All authors have read, edited, and approved the final manuscript.

## Data Availability

The datasets generated and analysed in the study are available in the Gene_Network_Project repository, https://github.com/ok-tsar/Gene_Network_Project.

## Acknowledgements

We would like to express our gratitude to the members of the Bioinformatics and Systems Medicine Laboratory (BSML), The Center for Precision Health, The University of Texas Health Science Center at Houston.

## Supplementary Information

**Additional file 1: Figure S1.** Overview of models for genotype‒phenotype relationships with the top 100 principal components. Three models—logistic regression (LOG), multilayer perceptron (MLP), and convolutional neural network (CNN)—each of which uses the top 100 kernel PCs from 15,132 genes.

**Additional file 2: Figure S2.** Evaluation of models for genotype‒phenotype discernment via 5-fold cross-validation. Detailed performance of the LOG, MLP, and CNN models. The data included average AUROC and AUPRC scores.

**Additional file 3: Figure S3.** Performance evaluation on test and unseen validation sets. LOG, MLP, and CNN models were assessed on a test set (GSE96058) and a validation set (GSE81538).

**Additional file 4: Figure S4.** Identifying optimal gene importance cutoffs in predictive models. Graphs showing AUROC and AUPRC scores for the top-ranked genes. The optimal cutoff for gene inclusion is indicated.

**Additional file 5: Figure S5.** Clustering performances of various clustering algorithms for all the models. Clustering recommendation and {selected parameter} for LOG: agglomerative clustering - {’n_clusters’: 5, ‘affinity’: ‘l1’, ‘linkage’: ‘average’}, MLP: agglomerative clustering - {’n_clusters’: 9, ‘affinity’: ‘euclidean’, ‘linkage’: ‘average’}, CNN: agglomerative clustering - {’n_clusters’: 10, ‘affinity’: ‘euclidean’, ‘linkage’: ‘complete’}.

**Additional file 6: Table S1.** Feature selection methodologies, their features, and pseudocode. The full code is available on the GitHub page.

**Additional file 7: Table S2.** Smart clustering: methodologies, search parameters, and features.

**Additional file 8: Table S3.** Literature review of critical genes identified by the LOG model. The critical genes are listed in descending order of importance with annotations on breast cancer, biomarkers, and therapeutic targets.

**Additional file 9: Table S4.** Literature review of critical genes identified by the MLP model. The critical genes are listed in descending order of importance with annotations on breast cancer, biomarkers, and therapeutic targets.

**Additional file 10: Table S5.** Literature review of critical genes identified by the CNN model. The critical genes are listed in descending order of importance with annotations on breast cancer, biomarkers, and therapeutic targets.

**Additional file 11: Table S6.** Model-derived and GeneMania network analyses across models. Locations of networks derived from LOG, MLP, and CNN models against GeneMania’s output, emphasizing similarities within networks. The table illustrates networks generated by our methodology for all models, ranked by in-network similarity.

