## Supplementary figures and images for "A Model-agnostic Computational Method for Discovering Gene–Phenotype Relationships and Inferring Gene Networks via *in silico* Gene Perturbation"

### Supplemental Figure 1

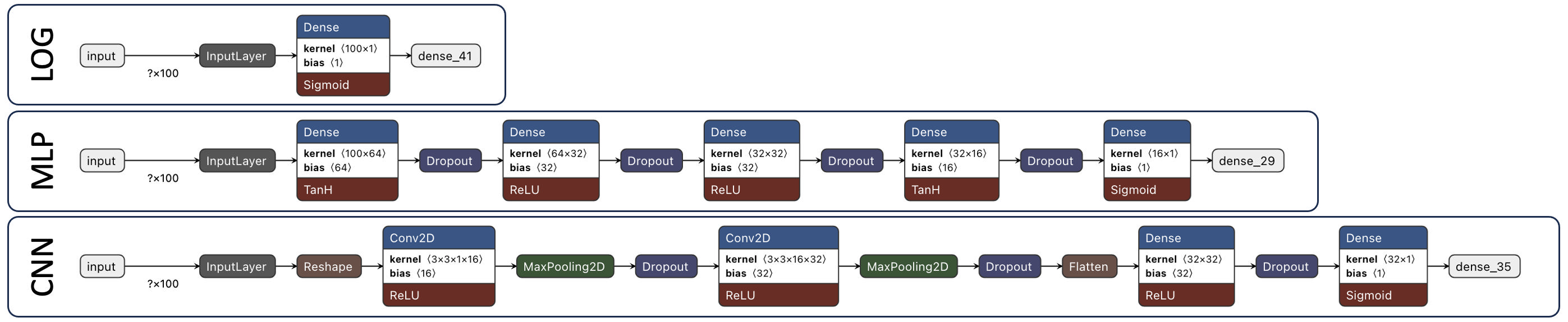

### Supplemental Figure 2

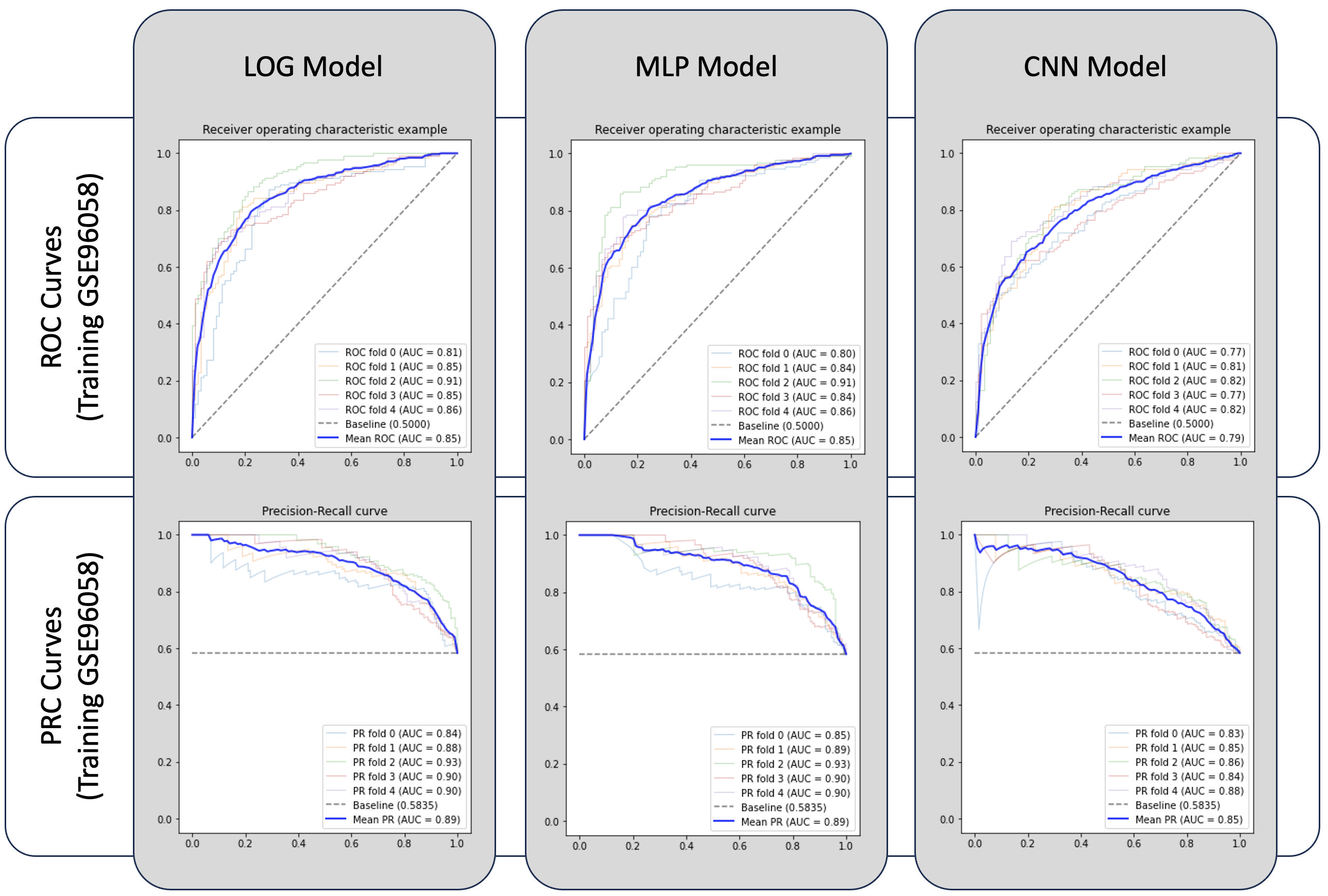

### Supplemental Figure 3

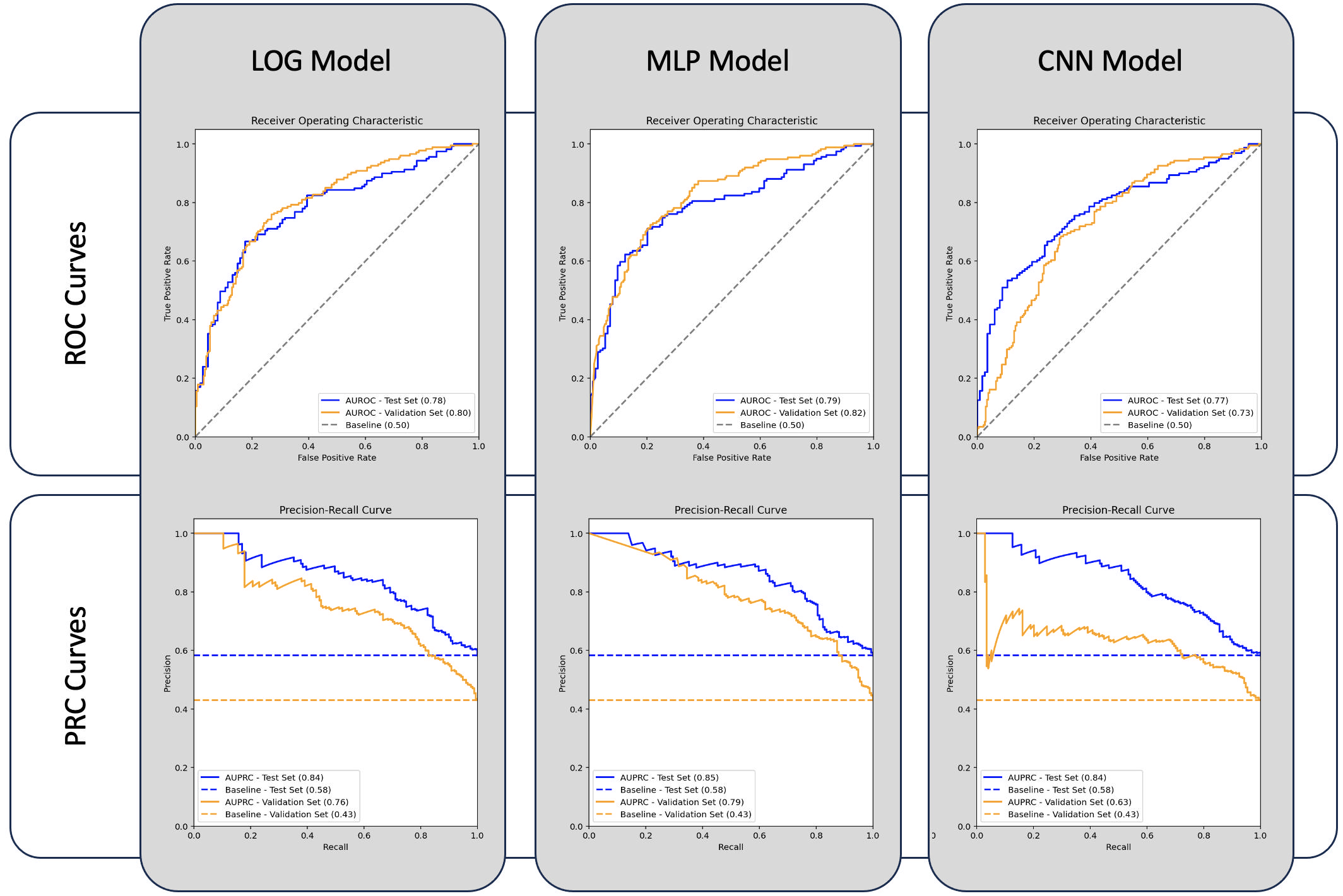

### Supplemental Figure 4

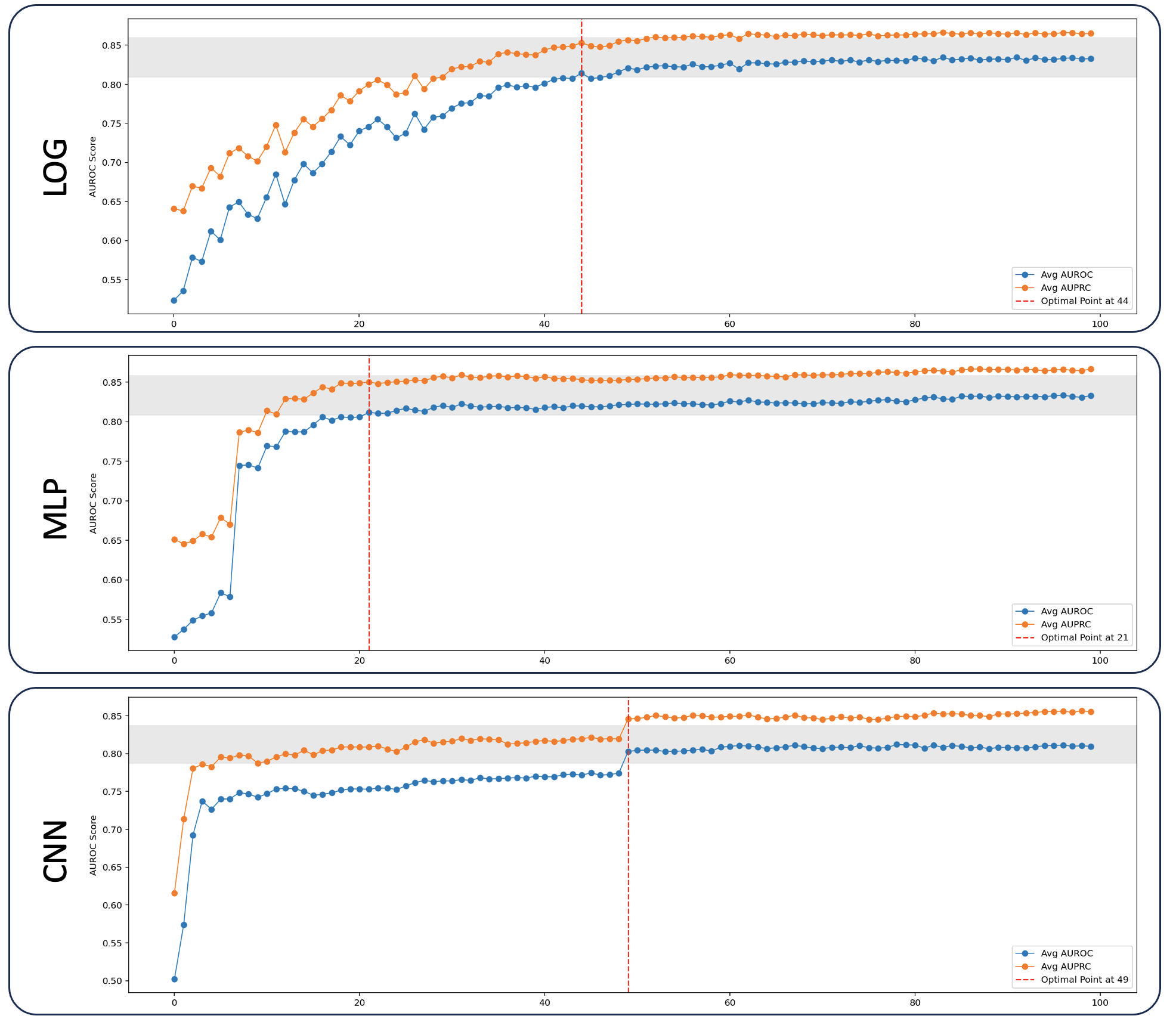

### Supplemental Figure 5

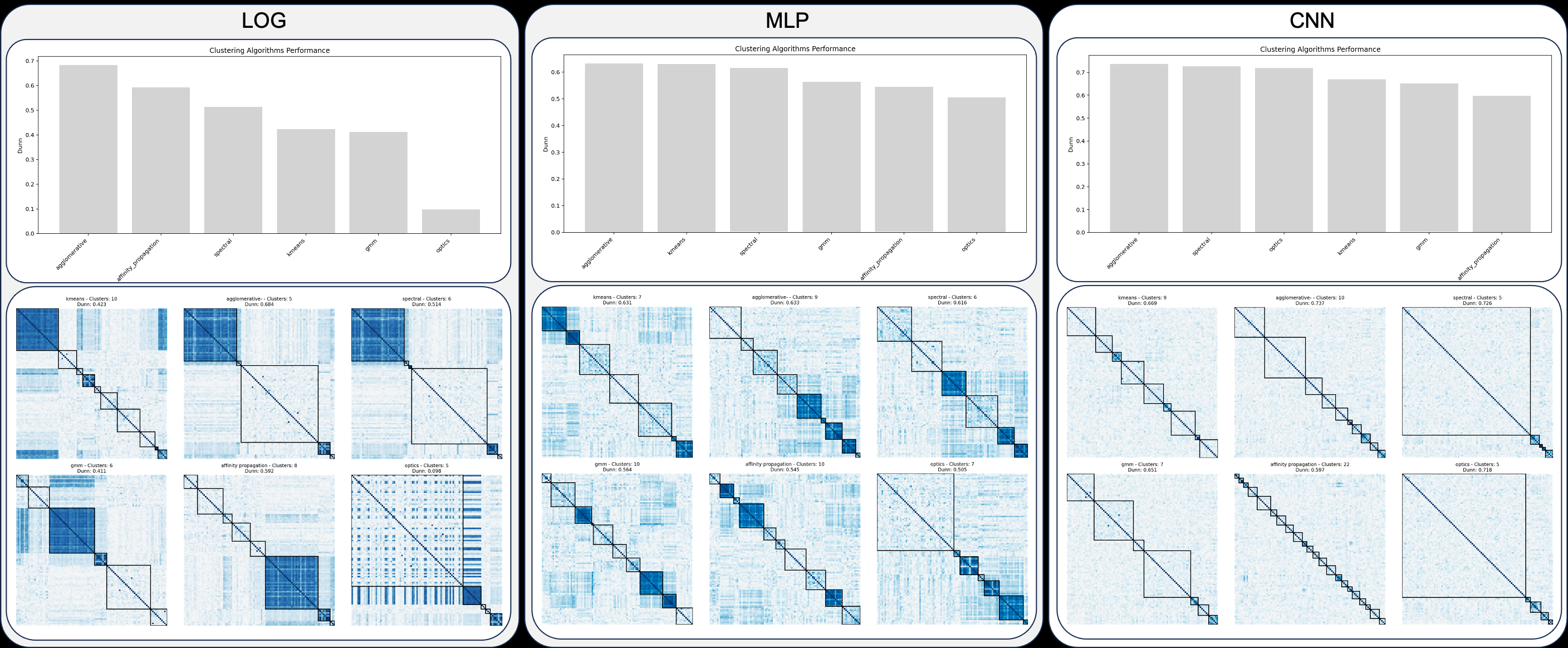
